# “Machine Learning–Based Prediction of Postoperative Refraction in Cataract Surgery: A Stacking Ensemble Approach”

**DOI:** 10.64898/2026.01.24.26344648

**Authors:** M. Gabar Zeyadi, Selcan Ipek-Ugay

## Abstract

**Purpose:** To develop and evaluate a stacking ensemble machine learning model for predicting postoperative spherical equivalent (SEQ) refraction after cataract surgery and to compare its performance with established intraocular lens power calculation formulas.

**Design:** Retrospective dataset study using a publicly available, de-identified dataset.

**Subjects:** A total of 1,710 eyes from 1,710 patients who underwent cataract surgery with monofocal intraocular lens implantation (Vivinex or SA60AT).

**Methods:** Following preprocessing and feature engineering, **we developed** a stacking ensemble architecture comprising three base learners - Multi-Layer Perceptron (MLP), Support Vector Regressor (SVR) with a radial basis function (RBF) kernel, and Spline Transformer with Linear Regression - and a Ridge Regressor meta-learner. We trained the model on 80% of the data (n=1,368) using 5-fold cross-validation and evaluated it on an independent 20% test set (n=342). We compared performance against six standard IOL formulas: Barrett Universal II, Kane, Haigis, Hoffer Q, Holladay 1, and SRK/T.

**Main Outcome Measures:** Mean absolute error (MAE), median absolute error (MedAE), root mean squared absolute error (RMSAE), standard deviation (SD), and percentage of eyes achieving absolute prediction error within ±0.25 D, ±0.50 D, ±0.75 D, and ±1.00 D.

**Results:** The stacking ensemble model demonstrated excellent predictive accuracy, achieving a Mean Absolute Error (MAE) of 0.271 D on the independent test set (n=342). The model achieved the lowest MAE among all evaluated methods and demonstrated statistically significant improvement over Barrett Universal II (MAE 0.317 D, p=0.004), Haigis, Holladay 1, SRK/T, and Hoffer Q (all p<0.001). The numerically lower MAE compared with the Kane formula (MAE 0.294 D, p=0.172) did not reach statistical significance in the primary analysis.

**Conclusion:** The stacking ensemble machine learning model achieves statistically significant improvement over five established IOL calculation formulas, demonstrating the potential of ensemble approaches in IOL power prediction. By leveraging algorithmic diversity and data-driven learning, this approach represents a promising advancement toward reducing refractive surprises and improving patient satisfaction in cataract surgery. External validation on independent datasets is warranted to assess generalizability.

## INTRODUCTION

Cataract surgery is among the most frequently performed surgical procedures globally, with modern objectives extending beyond visual restoration to achieving precise postoperative refractive outcomes.^1^ The accurate calculation of intraocular lens (IOL) power directly determines the postoperative spherical equivalent (SEQ) and patient visual quality of life.^2^ However, achieving this precision remains challenging, with “refractive surprises” - clinically significant deviations from intended outcomes - causing patient dissatisfaction and potentially requiring additional corrective procedures.^3^ The fundamental challenge lies in predicting the postoperative effective lens position (ELP), which clinicians cannot measure preoperatively and must estimate from biometric data.^4,5^

IOL calculation formulas have evolved from simple regression-based methods to sophisticated theoretical models.^6^ Third-generation formulas (SRK/T, Hoffer Q, Holladay 1) incorporated optical principles but relied on limited biometric variables.^7^ Fourth-generation formulas (Haigis, Barrett Universal II) integrated additional parameters to refine ELP predictions.^8,9^ Recently, artificial intelligence–enhanced formulas like Kane have emerged as clinical benchmarks, combining theoretical optics with machine learning to optimize predictions.^10,11^ Despite these advances, even state-of-the-art formulas achieve ±0.50 D accuracy in only 75%-90% of cases,^12^ and performance degrades in eyes with atypical biometry - each formula representing a single algorithmic hypothesis with an inherent performance ceiling.

Machine learning ensemble methods, which combine multiple diverse algorithms to achieve more robust predictions than any single model, offer a potential solution.^13,14^ The Nallasamy formula recently demonstrated proof-of-concept for stacking ensemble approaches in IOL calculation.^15^ However, the optimal stacking architecture -choice and diversity of base learners, selection of meta-learner, and their relative contributions- remains incompletely explored. This study aimed to develop and validate a stacking ensemble machine learning model for predicting postoperative SEQ in cataract surgery and to compare its performance against established IOL calculation formulas, including current clinical benchmarks. We hypothesized that: (**1**) a stacking ensemble with diverse base learners would outperform individual models and traditional formulas; (**2**) the ensemble would achieve accuracy comparable to current benchmarks (Kane, Barrett Universal II); and (**3**) algorithmic diversity would provide robust performance across different biometric ranges.

## METHODS

### Study Design and Dataset

We utilized a dataset of 1,710 eyes from 1,710 patients who underwent cataract surgery at the Augen- und Laserklinik Castrop Rauxel, Germany.

### Ethics

This study used publicly available, de-identified data released by the data provider (Figshare repository). No new data were collected for this analysis. The research adhered to the tenets of the Declaration of Helsinki. The Institutional Review Board of Berlin University of Applied Sciences determined that this study, utilizing publicly available de-identified data, did not constitute human subjects research and therefore did not require IRB approval. Informed consent was waived by the IRB as the study used publicly available, de-identified data.

The dataset consisted of two subsets based on the implanted monofocal IOL: 888 eyes received the Vivinex (Hoya Surgical Optics)^16^ and 822 eyes received the SA60AT (Alcon).^17^

### Patient Eligibility Criteria

The inclusion criteria required patients to have undergone successful cataract surgery, with no history of prior ophthalmic surgical interventions, no perioperative or postoperative complications, and no ocular pathologies that could influence biometric measurements, including corneal or retinal pathologies. An experienced optometrist measured the refractive power of the inserted lens and the postoperative refraction (sphere and cylinder) at least 4 weeks after surgery. The datasets included only data with a postoperative Snellen decimal visual acuity of 0.8 (20/25 Snellen lines) or higher to ensure reliable refraction measurements.

### Biometric Measurements

Preoperative biometry was performed using the IOLMaster 700 (Carl Zeiss Meditec, Jena, Germany). Initial parameters included axial length (AL), central corneal thickness (CCT), anterior chamber depth (ACD), aqueous depth (AQD), lens thickness (LT), corneal radii (R1, R2), keratometry (K1, K2), and white-to-white distance (WTW). Additionally, the refractive power of the inserted intraocular lens (IOL) and postoperative refraction measurements (SEQ) were recorded. (SEQ: spherical equivalent power of postoperative refraction).

### Data Preprocessing and Feature Engineering

Multicollinearity analysis using Variance Inflation Factor (VIF < 5 threshold):

- We observed high correlations between ACD and AQD. ACD was retained over AQD.
- We retained K1 and K2 (keratometry values) over R1 and R2 (corneal radii). Keratometry readings, measured in diopters, directly represent the refractive power of the cornea and are therefore more directly relevant to the task of predicting a refractive outcome.
- We excluded CCT due to low correlation with SEQ (Pearson r = 0.08, p = 0.31) and minimal contribution in recursive feature elimination.

**The final feature set comprised eight variables**: AL, ACD, LT, K1, K2, WTW, IOL power, and IOL type. The outcome variable (target) was SEQ. The clinical data are summarized in **Table 1**

**Table 1:** Summary of clinical data in the entire dataset (n = 1710)

| Parameters | Mean ± SD |
| --- | --- |
| Mean average K (D) | 43.70±1.54 |
| AL (mm) | 23.64±1.53 |
| ACD (mm) | 3.12±0.41 |
| LT (mm) | 4.62±0.45 |
| IOL power (D) | 21.64±4.30 |
| SEQ (D) | - 0.52±0.83 |

### Categorical Feature Encoding

We converted the categorical feature IOL type to numerical values using the LabelEncoder from the Scikit-learn library, which assigns integer values (0 and 1) to the two categories (Vivinex and SA60AT). For a binary categorical variable, this method is computationally efficient and functionally equivalent to one-hot encoding.

### Numerical Feature Scaling

The numerical features in the dataset possess different scales and units (e.g., AL in mm, K1 in D). Many machine learning algorithms, including the support vector regressor (SVR) and multi-layer perceptron (MLP) used in this study, are sensitive to the scale of input features. To ensure that all features contribute equitably to the model’s learning and to accelerate gradient-based optimization, we applied feature scaling. We used the StandardScaler from Scikit-learn to transform each numerical feature. This process standardizes the features by subtracting the mean and scaling to unit variance, resulting in a distribution with mean 0 and standard deviation 1. To prevent data leakage, the StandardScaler was fit exclusively to the training data, and the resulting scaler was subsequently used to transform both the training and test data.

### Data Partitioning Strategy

We split the dataset into a training set (80%; n=1,368) and an independent hold-out test set (20%; n=342). The test set was strictly held out and used only once for final, unbiased performance evaluation.

### Rationale for Ensemble Learning

To achieve the highest possible predictive accuracy, this study adopted a stacking ensemble machine learning framework. Ensemble methods combine the predictions of multiple models to produce a final prediction that is more accurate and robust than any of the individual constituent models. The core principle behind the stacking approach is that by combining the outputs of several different, well-performing models (known as base learners or Level-1 models), a second-level model (the meta-learner) can learn to optimally weigh these predictions, effectively correcting for the errors and biases of the individual models. This framework leverages the principle of model diversity; different algorithms, such as kernel methods, neural networks, and linear models, learn various aspects of the complex, non-linear relationships between the input features and the target variable. Stacking provides a principled, data-driven methodology for integrating diverse perspectives into a single, more powerful predictive system.

### Algorithm Selection

We selected constituent models for the stacking ensemble through a systematic, multi-stage process. A comprehensive screening of supervised learning regression algorithms was conducted, including tree-based methods (XGBoost, LightGBM, CatBoost, Random Forest),^18,19^ kernel methods (Kernel Ridge, SVR, Gaussian Process Regressor),^20^ linear models (Ridge, ElasticNet, Huber, SplineTransformer + Linear Regression),^21,22,23^ and neural networks (Multi-Layer Perceptron [MLP] Regressor).^24^ Following individual performance evaluations, the best-performing models were characterized by the lowest MAE. Overfitting was carefully checked, and the coefficients of the meta-learner were analyzed to gain insight into the relative weights and contributions assigned to each base model’s predictions.

### Stacking Ensemble Architecture

We developed a stacking ensemble regression framework to predict the postoperative spherical equivalent (SEQ) (**Figure 1**). The architecture consisted of two levels:

- **Level-0 (Base Learners):** Three diverse algorithms were selected to capture different data patterns: a Multi-Layer Perceptron (MLP) regressor for non-linear interactions,^24^ a Support Vector Regressor (SVR) with a radial basis function (RBF) kernel,^20^ and a SplineTransformer with Linear Regression. ^22,23^
- **Level-1 (Meta-Learner):** A Ridge Regressor ^21^ was employed to optimally weight the out-of-fold predictions from the base learners.

**Figure 1:**
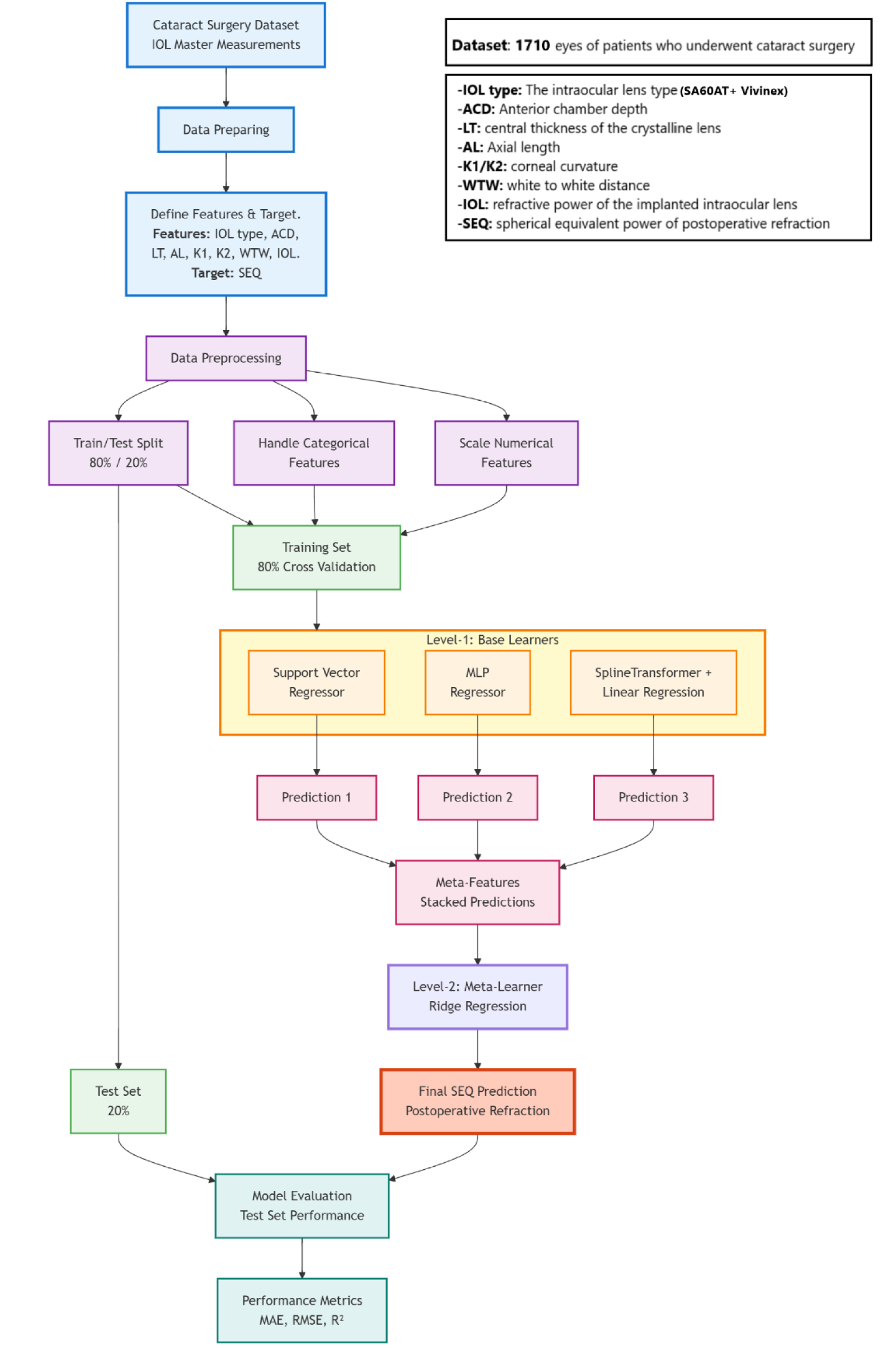
Stacking Ensemble Pipeline for SEQ Prediction

**We optimized** hyperparameters using Bayesian optimization (Optuna) ^25^ with 5-fold cross-validation on the training set, minimizing the Mean Absolute Error (MAE), preventing leakage and overfitting.

### Performance Evaluation and Statistical Analysis

The fundamental unit of evaluation is the prediction error, which was defined for each eye as:

### Error = actual postoperative refraction − predicted postoperative refraction

We evaluated the model’s predictive performance on the independent test set using Mean Absolute Error (MAE), Median Absolute Error (MedAE), and Root Mean Squared Absolute Error (RMSAE), Standard Deviation (SD), Mean Error (ME), R-squared (R^2^).

Clinical accuracy was quantified as the percentage of eyes with absolute prediction error within ±0.25 D, ±0.50 D, ±0.75 D, and ±1.00 D. A subgroup performance analysis was conducted based on axial length (AL).

We analyzed residuals for bias and distribution, normality tested using the Shapiro–Wilk test. The Agreement between predicted and actual SEQ was further assessed with Bland–Altman analysis (mean bias and 95% limits of agreement).

### Comparative Benchmark

We benchmarked the stacking ensemble model against six standard IOL calculation formulas: Barrett Universal II, Kane, Haigis, Hoffer Q, Holladay 1, and SRK/T. We assessed the statistical significance of differences in performance in the testing set using paired Wilcoxon signed-rank tests. Statistical significance was set at p < 0.05. For the primary analysis, Bonferroni correction was applied across all 21 pairwise comparisons among the six established formulas and the Stacking ML model. In addition, a targeted sensitivity analysis was performed by comparing the Stacking ML model with each of the six formulas, with Bonferroni correction applied for 6 comparisons, these results are presented in the Supplementary Materials. We used Bootstrap resampling with 1,000 iterations to compute 95% confidence intervals for all performance metrics.

We obtained Barrett Universal II and Kane predictions via their respective online calculators.^10,26^. For one eye in the test set, the Barrett Universal II prediction was not available at the time of initial data collection; the missing value was subsequently retrieved from the Barrett Universal II online calculator in March 2026, and the final comparative analysis was performed on the complete test set of n = 342.

We implemented Holladay 1,^27^ Hoffer Q, ^28,29^ SRK/T,^7,30^ and Haigis ^8,31^ in Python based on the published equations. For each implemented formula, we used IOLCon database constants for HOYA Vivinex and Alcon SA60AT IOL models as baseline values. These constants were further optimized by minimizing mean prediction error (zero-bias optimization) on the training set before applying them to the test set evaluation.

### Model Interpretability

SHapley Additive exPlanations (SHAP) analysis was employed for model interpretability. SHAP analysis quantified feature contributions to predictions, providing transparency and validating clinical plausibility.^32^

### Software and Tools Used

All computational tasks were performed using Python^33^ and its scientific computing ecosystem, including Scikit-learn,^34^ Pandas,^35^ NumPy,^36^ SciPy,^37^ Optuna,^25^ Tabulate,^38^ Matplotlib, Seaborn,^39^ and Google Colaboratory.^40^

## RESULTS

The stacking ensemble model achieved high predictive performance on the independent test set of 342 eyes. The stacking ensemble model achieved a Mean Absolute Error (MAE) of 0.271 D, a Mean Squared Error (MSE) of 0.121 D^2^, and a Standard Deviation (SD) of 0.348 D in the prediction errors. Furthermore, the model achieved a Coefficient of Determination (R^2^) of 0.808, indicating that it successfully explains approximately 80.8% of the variance in postoperative SEQ outcomes in the test set. The Mean Error (ME) of 0.013 D, close to zero, indicates a minimal systematic bias in the predictions. These performance metrics are summarized in **Table 2**, and **Figure 2** shows a scatter plot of Predicted vs. Actual Spherical Equivalent (SEQ).

**Figure 2:**
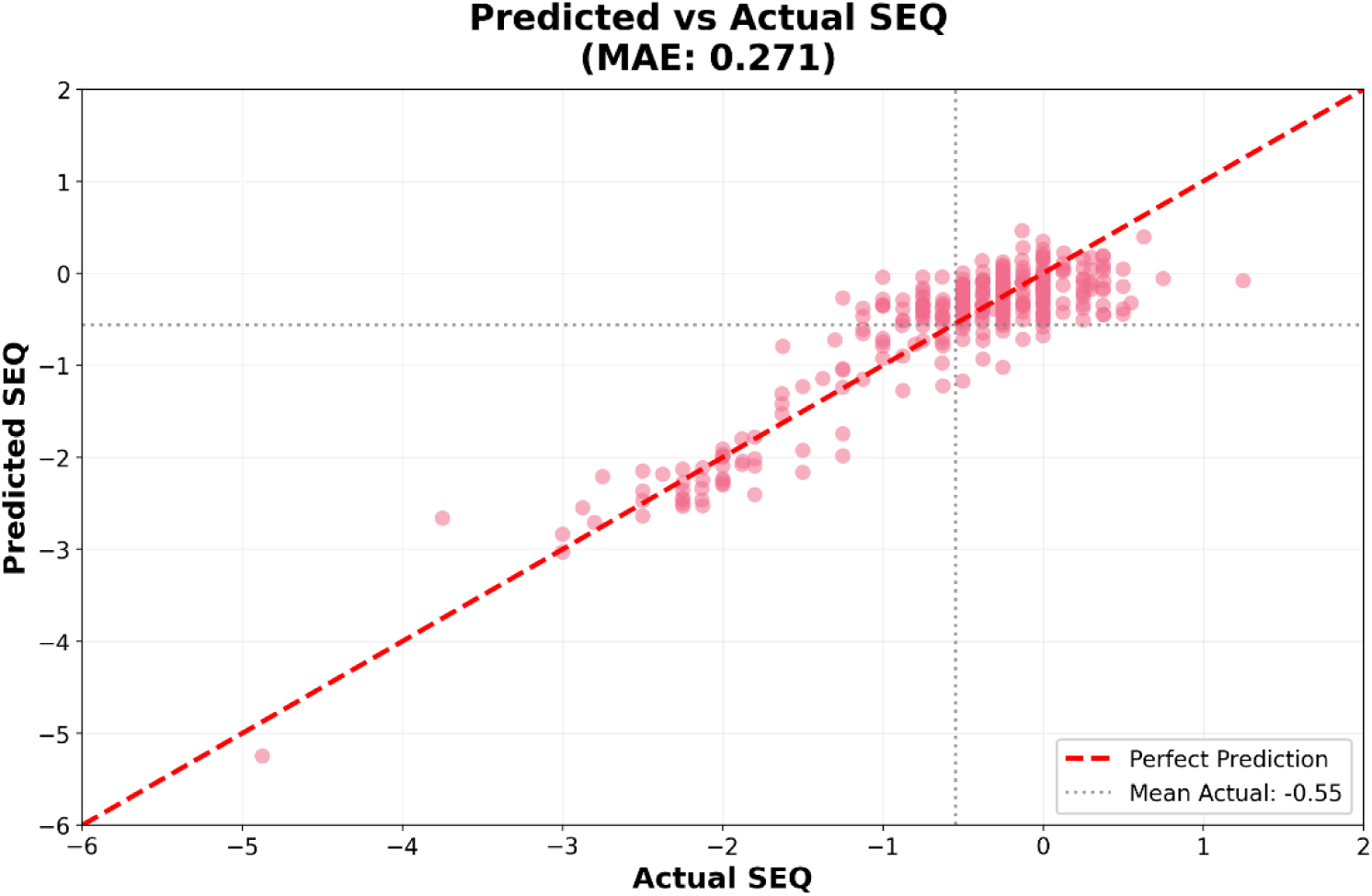
Predicted vs. Actual Spherical Equivalent (SEQ). This scatter plot illustrates the agreement between the predicted and actual postoperative SEQ values. The solid diagonal line represents the line of perfect prediction.

**Table 2:** Overall Performance Metrics of the Stacking Ensemble Model on the Independent Test Set (n=342 eyes).

| Metric | Value (D) |
| --- | --- |
| Mean Absolute Error (MAE) | 0.271 |
| Mean Squared Error (MSE) | 0.121 D<sup>2</sup> |
| R-squared (R<sup>2</sup>) | 0.808 |
| Mean Error (ME) | 0.013 |
| Standard Deviation of Prediction Error (SD) | 0.348 |

Training versus test performance showed excellent consistency (training MAE 0.266 D vs. test MAE 0.271 D), with a test-to-training MSE ratio of 1.039, indicating no overfitting. A summary of this comparative analysis is presented in **Table 3**.

**Table 3:**
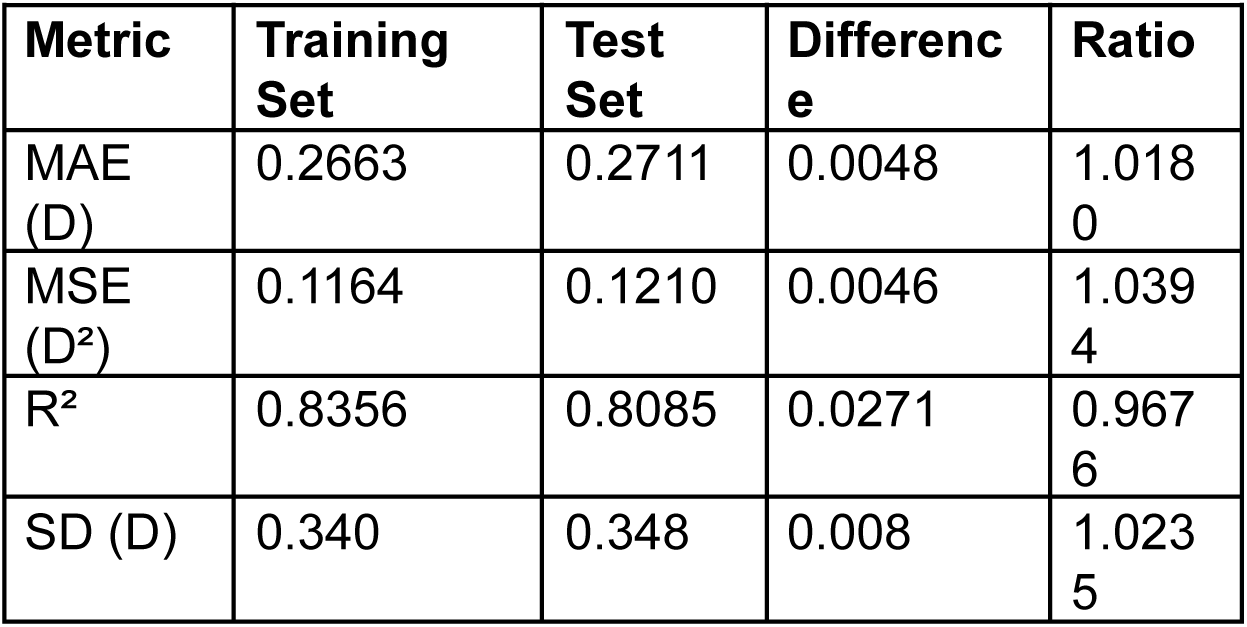
Training vs Test Performance Comparison of the Stacking Ensemble Model. MSE: Mean Squared Error, MAE: Mean Absolute Error, R^2^: = coefficient of determination, SD: Standard Deviation of the Prediction Error.

### Residual Analysis

Analysis of the prediction errors (residuals; Error = Actual SEQ – Predicted SEQ) on the test set demonstrated a mean of 0.013 D, indicating the absence of any significant systematic bias. The SD of the residuals was 0.348 D. A summary of this analysis is provided in **Table 4** and **Figure 3** shows a scatter plot of residuals vs. actual SEQ.

**Figure 3:**
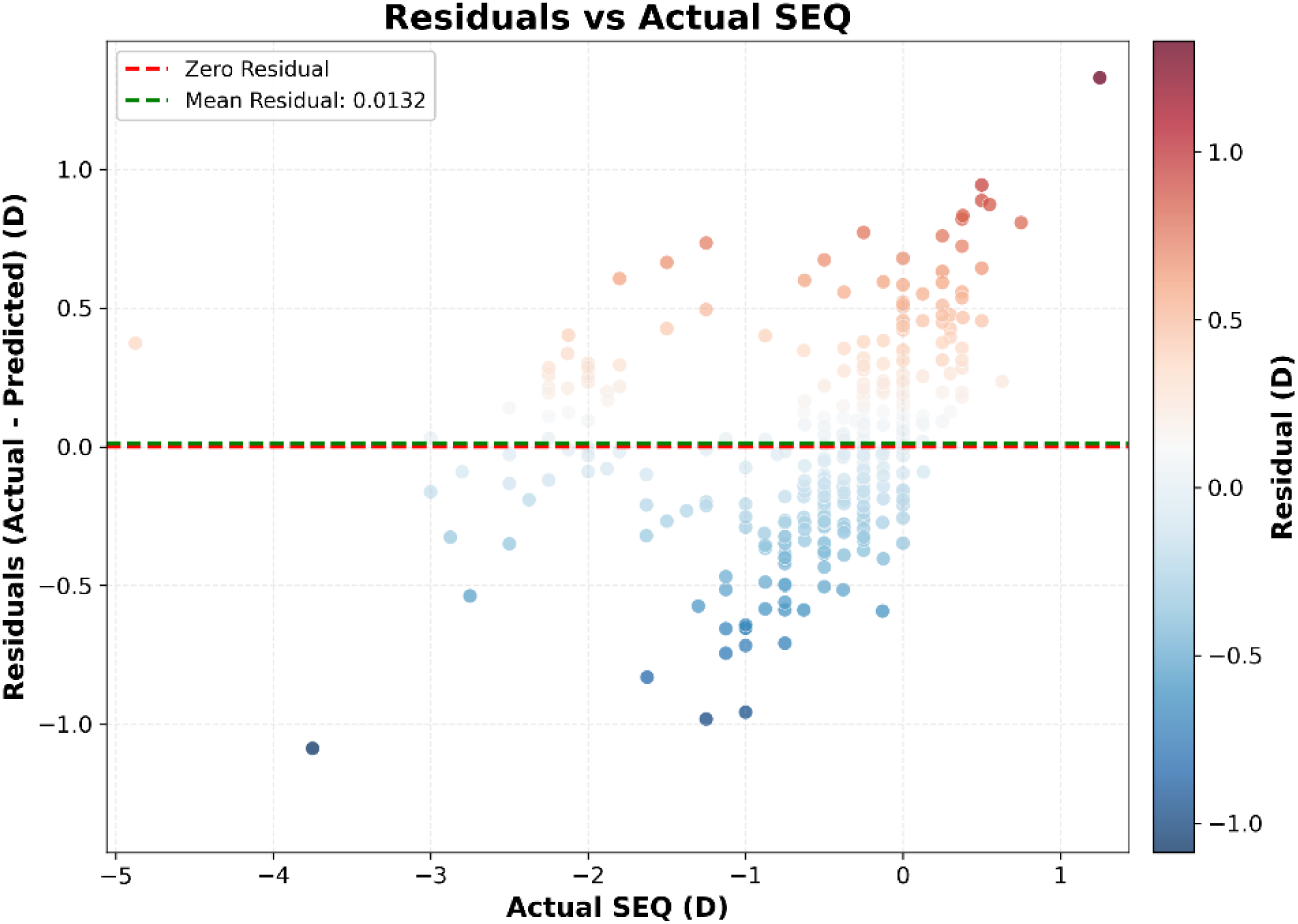
Residuals versus actual postoperative spherical equivalent (SEQ).

**Table 4:** Residual Analysis of the Stacking Ensemble Model on the Independent Test Set. LOA = limits of agreement; CI = confidence interval.

| Statistic | Value (D) |
| --- | --- |
| Mean Error (ME / Bias) | 0.013491 |
| Standard Deviation (SD) | 0.348 |
| Min | -1.087473 |
| Max | 1.330069 |
| Range | 2.417542 |
| 95% Limits of Agreement (LOA) | [-0.669, 0.696] |
| 95% CI for Mean Signed Error | [-0.023, 0.050] |

### Precision and Limits of Agreement

The SD of the prediction error was 0.348 D. Bland-Altman analysis established the 95% LOA between the predicted and actual SEQ at [−0.669 D, 0.696 D]. The 95% CI for the mean signed error ranged from −0.023 D to 0.050 D, confirming that the systematic bias is close to zero. **Figure 4** shows the Bland-Altman analysis.

**Figure 4.**
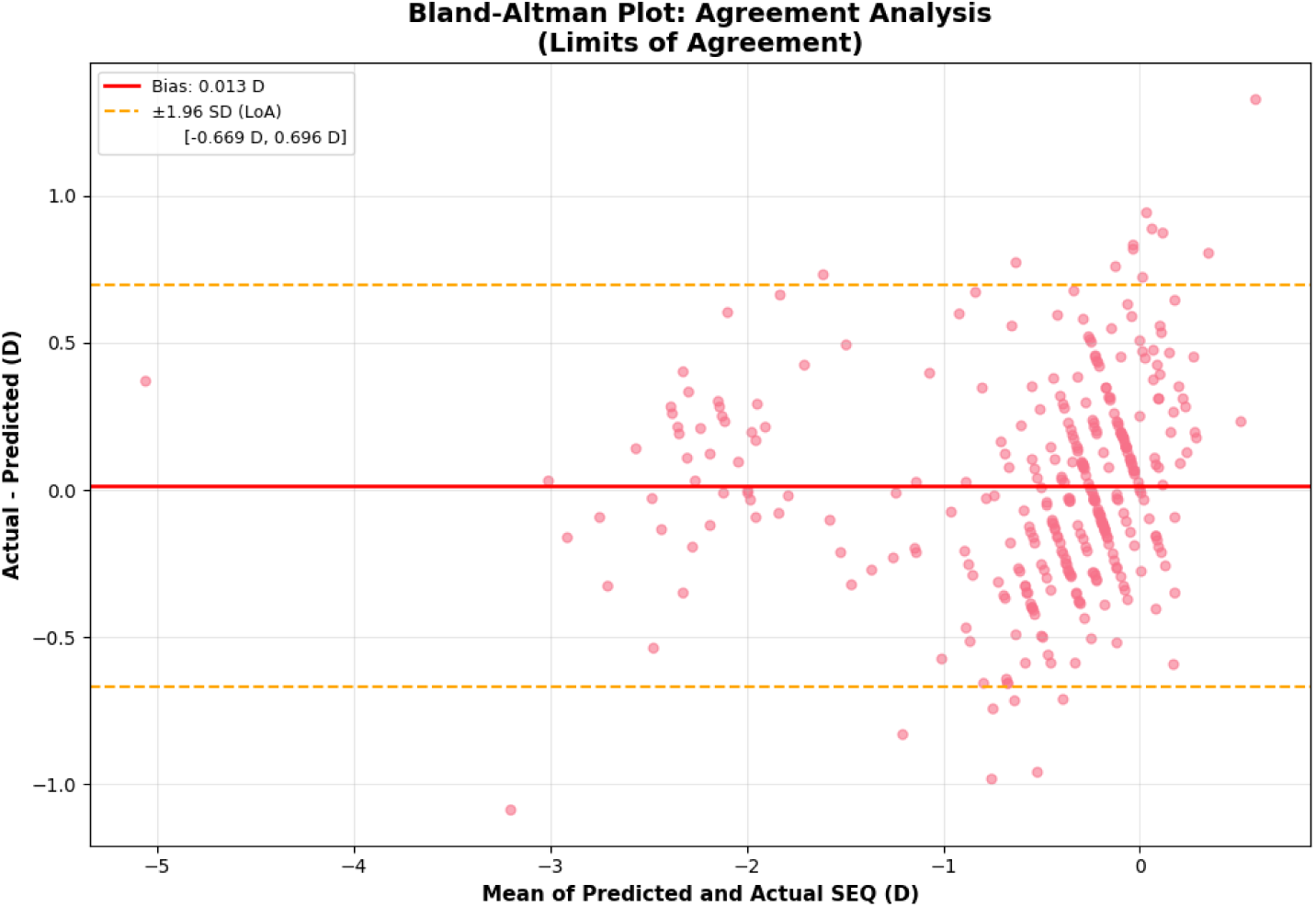
Bland–Altman Analysis. Mean signed error (bias) was 0.013 D, with 95% limits of agreement from −0.669 D to 0.696 D (bias ± 1.96 SD).

### Comparative Benchmark Against Standard IOL Formulas

We benchmarked the stacking ensemble model against six standard IOL calculation formulas on the independent test set of 342 eyes. The statistical prediction error metrics (MAE, MedAE, RMSAE, and SD) with bootstrap 95% confidence intervals are presented in **Table 5** and visualized in **Figure 5**. The stacking model achieved the lowest MAE (0.271 D [95% CI: 0.248–0.295] ) among all evaluated methods, followed by Kane (MAE 0.294 D [95% CI: 0.270–0.319], p=0.172 ), Haigis (0.315 D [95% CI: 0.289–0.341], p<0.001), and Barrett Universal II (0.317 D [95% CI: 0.287–0.350], p=0.004).

**Figure 5:**
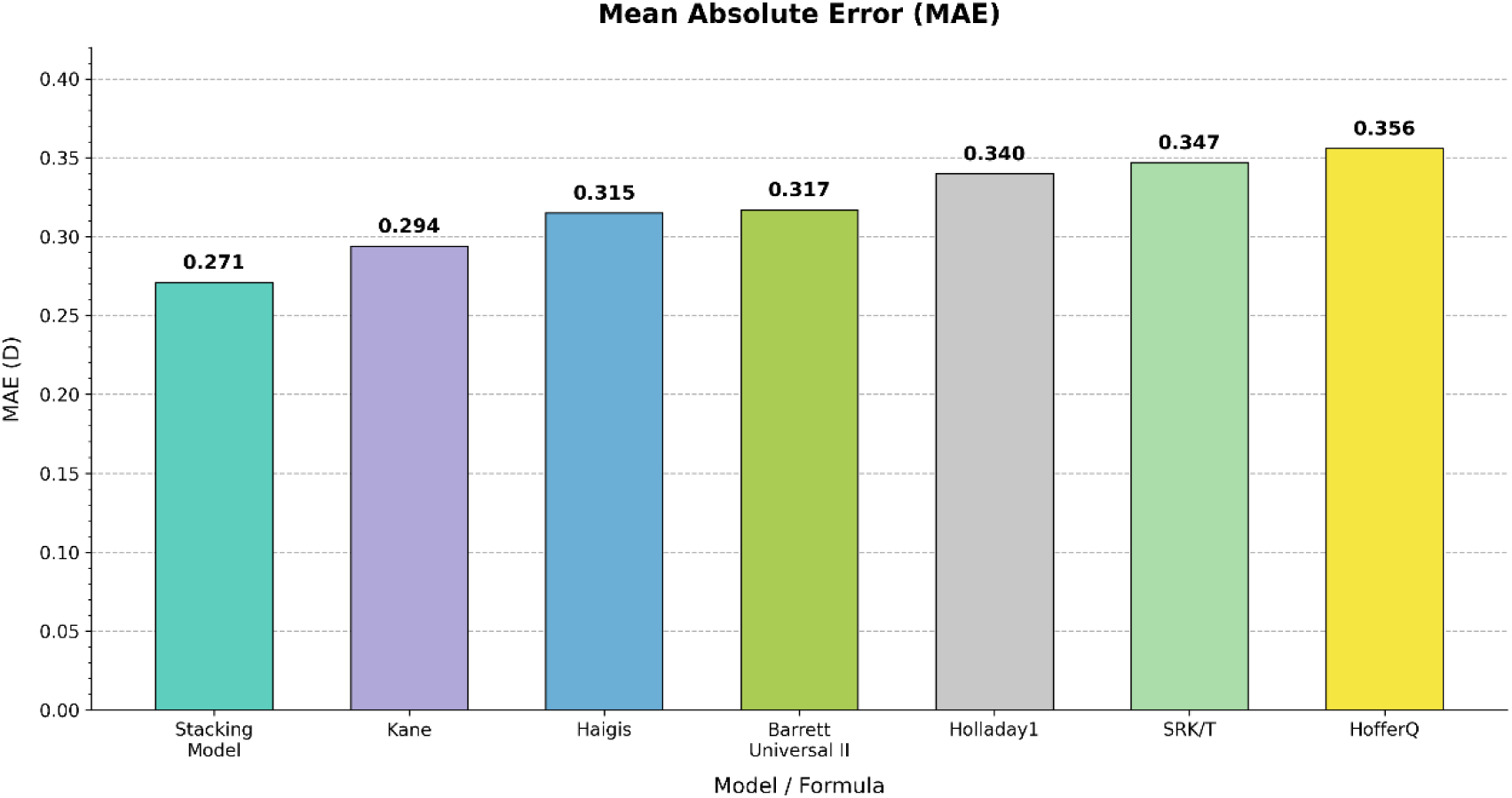
Prediction Performance for the stacking model and the six standard formulas

**Table 5:** Prediction Performance summary in the testing set (n=342 eyes). MAE: Mean Absolute Error with 95% confidence intervals computed via bootstrap resampling (1,000 iterations), MedAE: Median Absolute Error, RMSAE: Root Mean Squared Absolute Error, SD: Standard Deviation of the Prediction Error. P-values were derived from paired Wilcoxon signed-rank tests with Bonferroni correction applied across all 21 pairwise comparisons among the six established formulas and the Stacking ML model.

| Model/Formula | MAE [95% CI] | MedAE | RMSAE | SD | P-Value |
| --- | --- | --- | --- | --- | --- |
| Stacking Model | 0.271<br>[0.248-0.295] | 0.215 | 0.348 | 0.348 | -- |
| Kane | 0.294<br>[0.270–0.319] | 0.230 | 0.377 | 0.364 | 0.172 |
| Haigis | 0.315<br>[0.289–0.341] | 0.253 | 0.402 | 0.402 | < 0.001 |
| Barrett Universal II | 0.317<br>[0.287–0.350] | 0.250 | 0.431 | 0.422 | 0.004 |
| Holladay1 | 0.340<br>[0.310–0.370] | 0.272 | 0.443 | 0.444 | < 0.001 |
| SRK/T | 0.347<br>[0.315–0.381] | 0.270 | 0.458 | 0.458 | < 0.001 |
| HofferQ | 0.356<br>[0.325–0.387] | 0.288 | 0.460 | 0.461 | < 0.001 |

### Comparison of Clinical Accuracy Thresholds

We compared the clinical efficacy of each method by calculating the percentage of eyes achieving an absolute prediction error within the four standard clinical thresholds. As shown in **Table 6** and visualized in **Figure 6**, the stacking model achieved the highest success rate among all methods. Within ±0.50 D, the stacking model correctly predicted 85.1% of cases, compared with 82.5% for Kane and 81.8% for Barrett Universal II. At the ±0.25 D threshold, the stacking model achieved 54.7%, versus 53.8% for Kane and 51.3% for Barrett Universal II.

**Figure 6:**
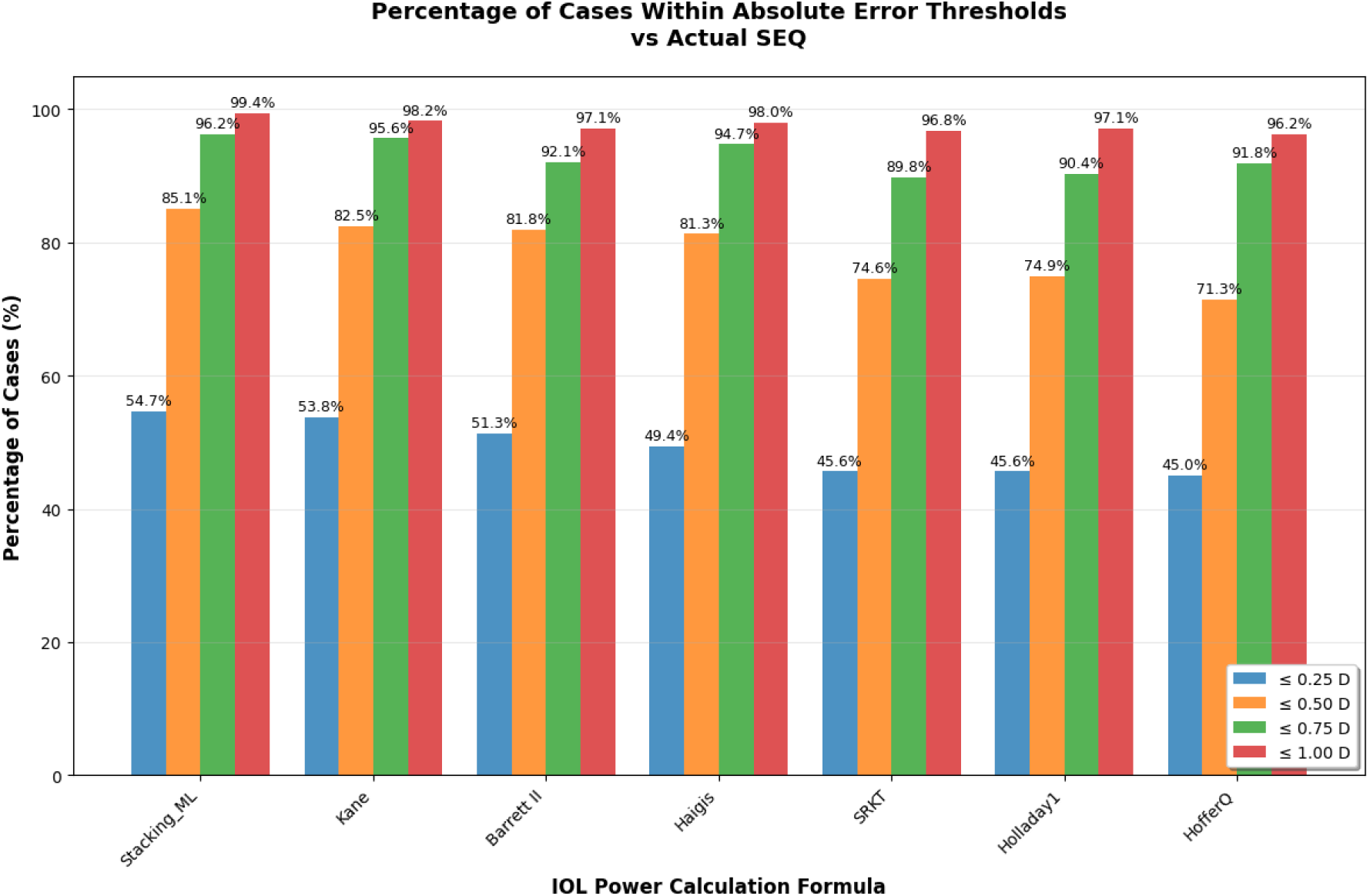
The percentage of Absolute Prediction Errors within Clinically Significant Thresholds (±0.25 D, ±0.50 D, ±0.75 D, ±1.00 D) for each formula, calculated based on the results in the testing dataset.

**Table 6:** The percentage of Absolute Prediction Errors within Clinically Significant Thresholds for each formula, calculated based on the results in the testing dataset.

| Model/ Formula | $\pm 0.25$ D | $\pm 0.50$ D | $\pm 0.75$ D | $\pm 1.00$ D |
| --- | --- | --- | --- | --- |
| Stacking Model | 54.7 % | 85.1 % | 96.2 % | 99.4 % |
| Kane Formula | 53.8 % | 82.5 % | 95.6 % | 98.2 % |
| Barrett Universal II | 51.3 % | 81.8 % | 92.1 % | 97.1 % |
| Haigis Formula | 49.4 % | 81.3 % | 94.7 % | 98.0 % |
| Holladay 1 Formula | 45.6 % | 74.9 % | 90.4 % | 97.1 % |
| SRK/T Formula | 45.6 % | 74.6 % | 89.8 % | 96.8 % |
| Hoffer Q Formula | 45.0 % | 71.3 % | 91.8 % | 96.2 % |

### Subgroup Analysis by Axial Length

Performance varied across eye sizes. As shown in **Table 7** and visualized in **Figures 7 and 8**, the stacking model demonstrated the lowest MAE in the medium AL group and the short AL group. In the short AL group (<22.0 mm), the stacking model’s MAE of 0.386 D was lower than Kane’s (0.397 D). In the long AL group (>26.0 mm), Kane achieved an MAE of 0.205 D, compared with 0.231 D for the stacking model.

**Figure 7:**
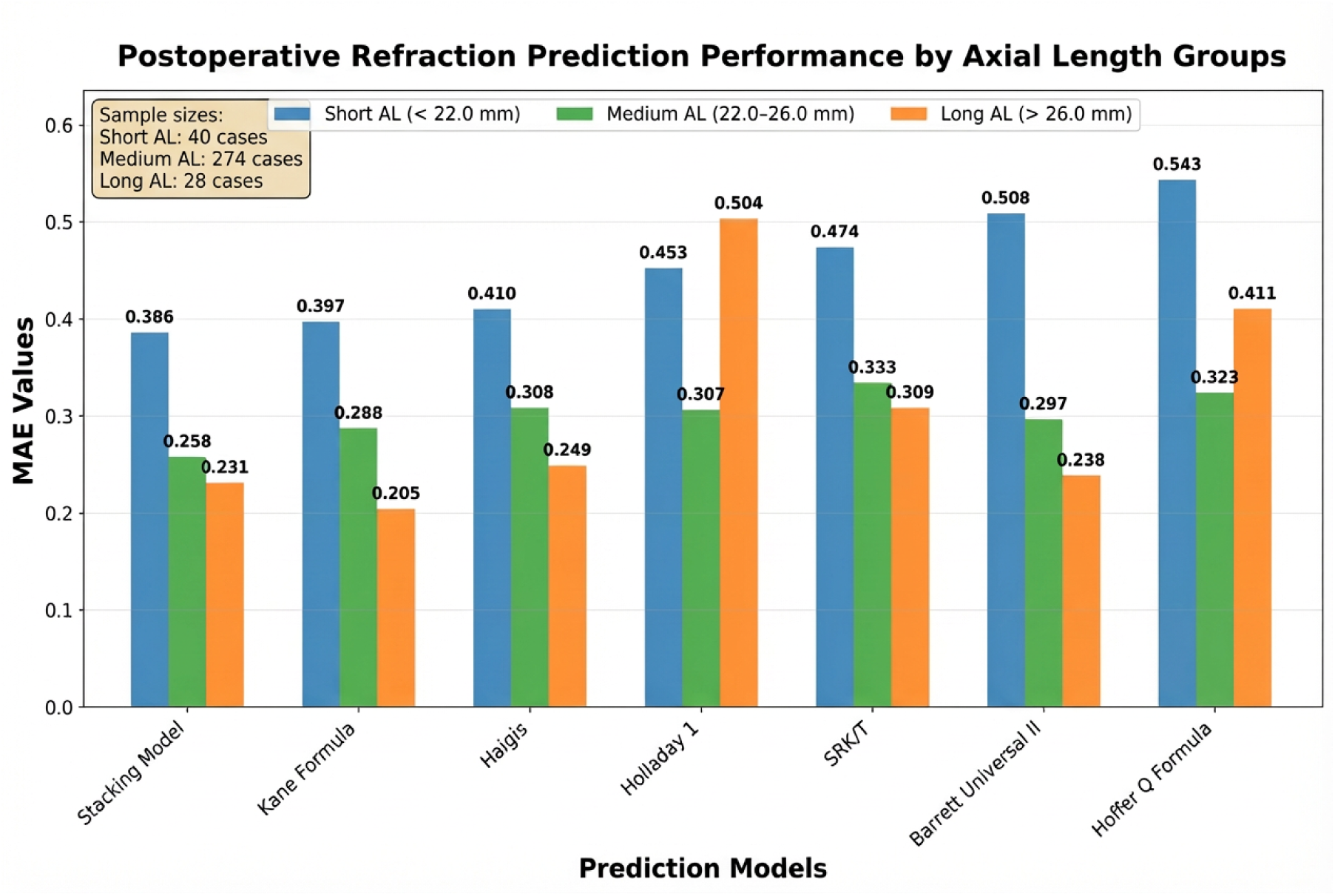
Prediction Performance Analysis by Axial Length MAE: Mean Absolute Error. AL: Axial length.

**Figure 8:**
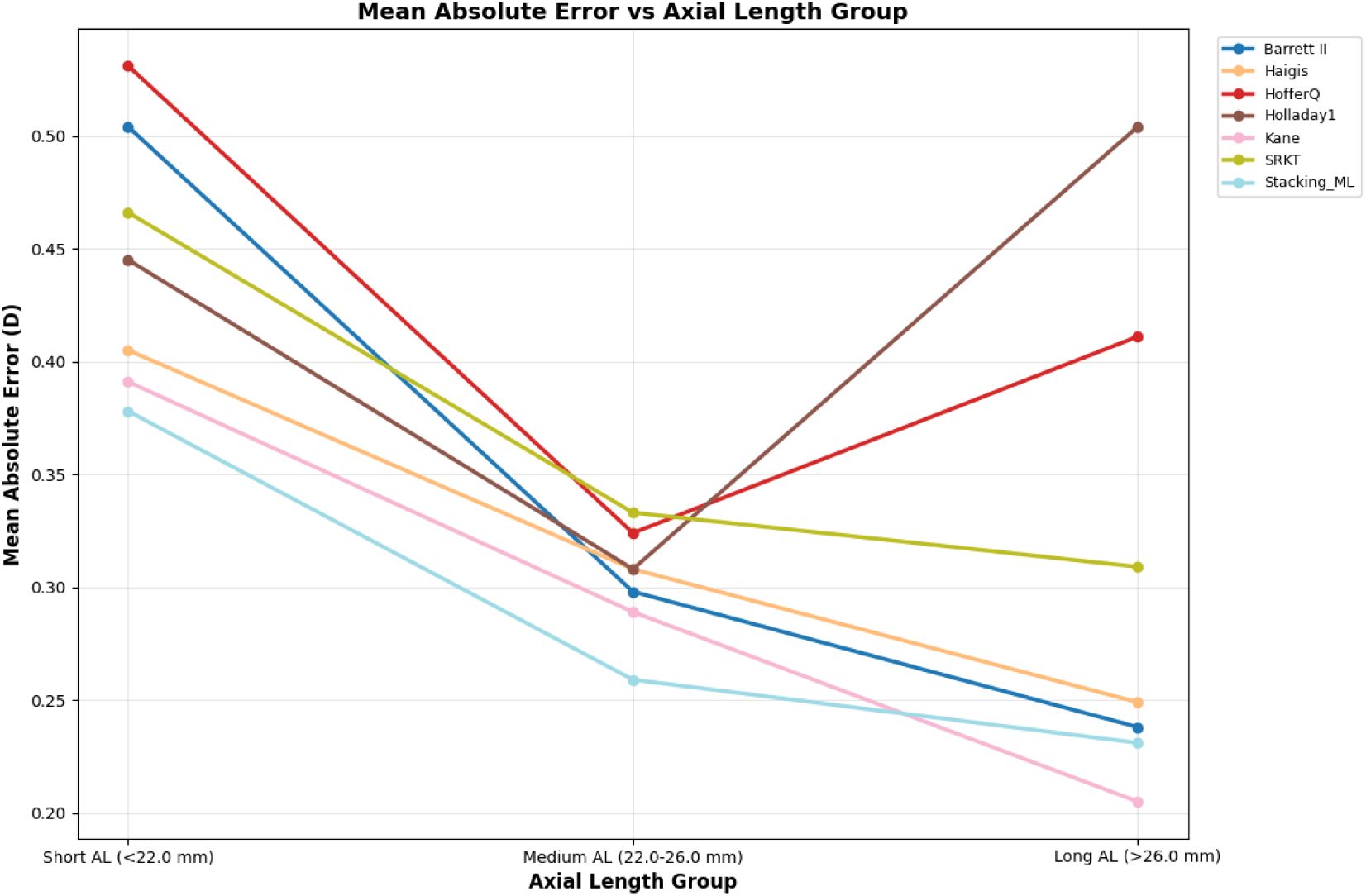
Mean Absolute Error vs Axial Length

**Table 7:** Prediction Performance Analysis by Axial Length. MAE: Mean Absolute Error, MedAE: Median Absolute Error, RMSAE: Root Mean Squared Absolute Error, SD: Standard Deviation of the Prediction Error, N: Number of cases.

| **Axial Length Group** | **Model / Formula** | **N** | **MAE** | **MedAE** | **RMSAE** | **SD** |
| --- | --- | --- | --- | --- | --- | --- |
| **Short AL (<22.0 mm)** | **Stacking Model** | 40 | **0.386** | **0.312** | **0.467** | **0.466** |
|  | Kane Formula | 40 | 0.397 | 0.350 | 0.485 | 0.478 |
|  | Haigis | 40 | 0.410 | 0.352 | 0.526 | 0.532 |
|  | Holladay 1 | 40 | 0.453 | 0.381 | 0.573 | 0.566 |
|  | SRK/T | 40 | 0.474 | 0.415 | 0.618 | 0.625 |
|  | Barrett Universal II | 40 | 0.508 | 0.398 | 0.687 | 0.695 |
|  | Hoffer Q Formula | 40 | 0.543 | 0.500 | 0.674 | 0.587 |
| **Medium AL (22.0–26.0 mm)** | **Stacking Model** | 274 | **0.258** | **0.199** | **0.333** | **0.334** |
|  | Kane | 274 | 0.288 | 0.223 | 0.369 | 0.353 |
|  | Barrett Universal II | 274 | 0.297 | 0.233 | 0.392 | 0.379 |
|  | Holladay 1 | 274 | 0.307 | 0.251 | 0.403 | 0.400 |
|  | Haigis | 274 | 0.308 | 0.248 | 0.389 | 0.388 |
|  | Hoffer Q | 274 | 0.323 | 0.257 | 0.419 | 0.419 |
|  | SRK/T | 274 | 0.333 | 0.261 | 0.436 | 0.433 |
| **Long AL (>26.0 mm)** | **Kane** | 28 | **0.205** | **0.160** | **0.253** | **0.257** |
|  | Stacking Model | 28 | 0.231 | 0.216 | 0.283 | 0.274 |
|  | Barrett Universal II | 28 | 0.238 | 0.240 | 0.302 | 0.272 |
|  | Haigis | 28 | 0.249 | 0.212 | 0.303 | 0.293 |
|  | SRK/T | 28 | 0.309 | 0.276 | 0.389 | 0.310 |
|  | Hoffer Q | 28 | 0.411 | 0.435 | 0.474 | 0.304 |
|  | Holladay 1 | 28 | 0.504 | 0.527 | 0.586 | 0.320 |

### Model Interpretability

SHAP analysis (**Figure 9**) identified Axial Length and IOL Power as the dominant predictive features, followed by keratometry (K1, K2), lens thickness, and anterior chamber depth. This feature importance hierarchy aligns with established optical theory, where AL and IOL power are the primary determinants of postoperative refraction. This feature prioritization aligns with established optical principles, confirming the clinical plausibility of the model’s decision-making process.

**Figure 9:**
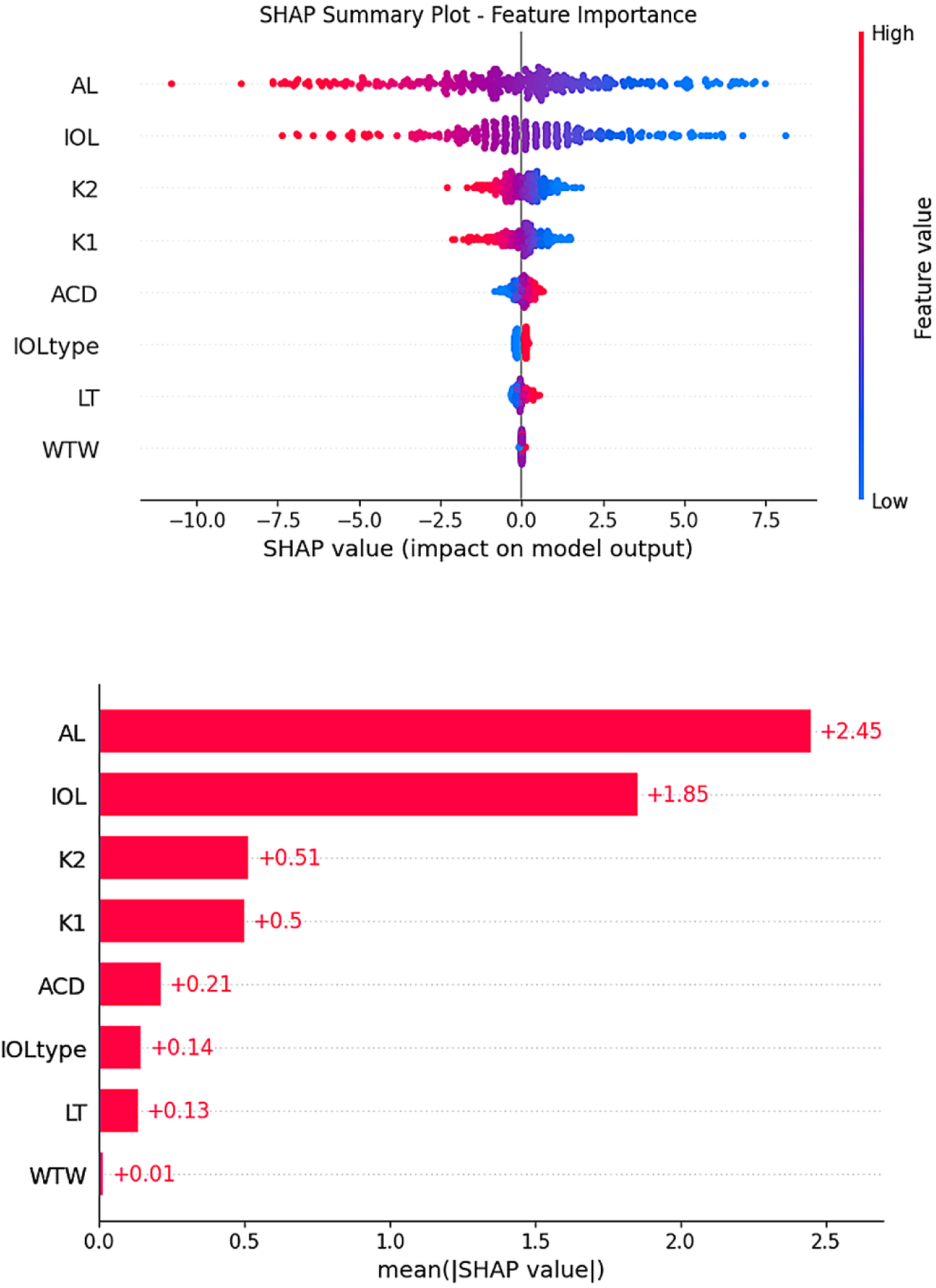
SHapley Additive exPlanations (SHAP) analysis showing the features influencing the stacking model’s predictions.

## DISCUSSION

### Principal Findings

This study demonstrates that the stacking ensemble machine learning model achieves statistically significant improvement over five of six evaluated IOL calculation formulas: Barrett Universal II (p=0.004), Haigis, Holladay 1, SRK/T, and Hoffer Q (all p<0.001). The model achieved numerically lower MAE than the Kane formula (0.271 D vs. 0.294 D), though this difference did not reach statistical significance in the primary analysis (p=0.172).

The stacking model achieved a Mean Absolute Error (MAE) of 0.271 D **(Table 5)** and predicted 85.1% of eyes within ±0.50 D of the target refraction **(Table 6)** outperforming Kane formula (MAE 0.294 D; 82.5% within ±0.50 D), Barrett Universal II (MAE 0.317 D; 81.8% within ±0.50 D) and all other evaluated formulas.

### Model Architecture and Performance

The performance of the ensemble architecture is attributable to algorithmic diversity (**Figure 1**). By combining a neural network (MLP), a kernel-based method (SVR), and a spline-based linear model, the meta-learner could leverage the unique strengths of each. The MLP captures complex non-linear feature interactions, the SVR provides robustness to outliers with its epsilon-insensitive loss function, and the SplineTransformer offers flexible piecewise polynomial modelling. The Ridge meta-learner intelligently weights the predictions demonstrating data-driven synthesis that mitigates individual model weaknesses.

The model’s generalization capability was rigorously validated. Minimal training-test performance discrepancy (MAE 0.266 D vs. 0.271 D) (**Table 3**), close cross-validation alignment, near-zero mean error, and tight Bland-Altman (**Figure 4**) limits of agreement collectively confirm the model learned true underlying patterns rather than overfitting training data.

### Clinical Significance and Comparison with Current Standards

The 85.1% success rate at ±0.50 D represents excellent performance comparable to leading formulas. Recent literature confirms that 80-85% accuracy at this threshold represents excellent performance. ^12^

### Model Interpretability and Clinical Trust

SHAP analysis (**Figure 9**) addressed the critical “black box” concern inherent in complex machine learning models. The identification of AL and IOL power as dominant features, followed by keratometry values, precisely mirrors the theoretical foundation of IOL calculation. This alignment with established ophthalmological principles provides evidence that the model learns clinically relevant, causal relationships rather than spurious correlations. Such interpretability is essential for clinical adoption and regulatory acceptance.

### Study Strengths

This study employed methodological rigor across multiple dimensions. The large, carefully curated dataset (1,710 eyes) with high-quality biometry provided substantial statistical power. Algorithmic diversity in base learner selection maximized ensemble capability. The benchmarking strategy ensured fair comparison by evaluating all methods on identical test data. Finally, proactive SHAP analysis addressed interpretability, a sophisticated and vital component of modern clinical AI research ^32^.

### Limitations

Some limitations merit consideration. First, single-center data origin may limit external generalizability. Model performance may differ with other biometry devices, surgical techniques, or patient demographics, necessitating multi-center validation. Second, training exclusively on two monofocal IOL models (Vivinex and SA60AT) means generalizability to other IOL designs, particularly multifocal and Toric lenses, remains untested. Third, sex and gender dimensions were not analyzed in this study. Fourth, strict exclusion criteria removed patients with prior corneal refractive surgery (e.g., post-LASIK), significant ocular pathologies, or perioperative complications. While appropriate for foundational model development, performance in these complex cases which represent significant clinical challenges remains unproven. The model is currently validated for uncomplicated eyes only. The model was trained exclusively on data from IOLMaster 700 biometry; generalizability to other devices (e.g., IOLMaster 500, Lenstar, OA-2000) remains unvalidated. Additionally, the geographic and ethnic homogeneity of the single-center German dataset may limit applicability to diverse global populations. Fifth, while our model demonstrated numerically lower MAE than the Kane formula (0.271 D vs. 0.294 D), this difference did not achieve statistical significance (p=0.172). This may reflect limited statistical power due to the moderate test set size (n=342). Larger multi-center validation studies are needed to definitively establish relative performance against leading formulas. Sixth, our model achieved numerically lower MAE (0.271 D) than the Nallasamy stacking ensemble (0.312 D)^15^ but, unlike Nallasamy, did not achieve statistical significance against Kane. This discrepancy likely reflects differences in sample size (n=342 vs ∼1,000 eyes), population characteristics, or dataset-specific Kane formula performance variance. Multi-center validation with larger cohorts is essential to definitively establish comparative performance.

### Clinical Implications and Implementation

The demonstrated superior performance suggests strong potential for clinical integration as a decision-support tool. When existing formulas provide divergent predictions, the model’s output could guide final IOL power selection. The ultimate goal is consistently reducing refractive surprises unexpected hyperopic or myopic outcomes that compromise visual quality and necessitate secondary procedures.

### Future Research Directions

Multi-center external validation on large, heterogeneous datasets from multiple surgeons and biometry devices is essential. Model expansion should include diverse IOL designs and types (Multifocal, Toric). A dedicated research track should develop specialized models for post-refractive surgery eyes and other challenging scenarios.

## CONCLUSION

The stacking ensemble machine learning model achieves statistically significant improvement over five established IOL calculation formulas, demonstrating the potential of ensemble approaches in IOL power prediction. By leveraging algorithmic diversity and data-driven learning, this approach represents a promising advancement toward reducing refractive surprises and improving patient satisfaction in cataract surgery. External validation on independent datasets is warranted to assess generalizability.

## ACKNOWLEDGMENTS

The authors acknowledge Prof. Dr. Achim Langenbucher (Institute of Experimental Ophthalmology, Saarland University, Germany) for making the dataset publicly available, and the Augen- und Laserklinik Castrop-Rauxel, Germany, for the original data collection.

## Data Availability

The dataset used in this study is publicly available through the Figshare repository: https://figshare.com/authors/Achim_Langenbucher/12446667.

Preprocessed data are available from the corresponding author upon reasonable request.

## Code Availability

The Python implementation of the stacking ensemble model, all analysis scripts, and a Jupyter notebook demonstrating the complete pipeline are publicly available via GitHub (https://github.com/Gabar0/Stacking-Ensemble-SEQ-Prediction) and archived on Zenodo (https://doi.org/10.5281/zenodo.19443026).

